# Effect of relaxation interventions in pregnant women on maternal and neonatal outcomes: A systematic review and meta-analysis

**DOI:** 10.1101/2022.11.17.22282468

**Authors:** Mubarek Abera, Charlotte Hanlon, Beniam Daniel, Markos Tesfaye, Abdulhalik Workicho, Tsinuel Grima, Wibaek Rasmus, Gregers Andersen, Mary Fewtrell, Suzanne Filteau, Jonathan C Wells

## Abstract

**Background:** Maternal stress during pregnancy has been associated with adverse pregnancy and birth outcomes. Aiming to reduce maternal stress and to improve pregnancy and birth outcomes, different relaxation interventions have been tested during pregnancy. This systematic review and meta-analysis was conducted on studies that have tested relaxation interventions to improve maternal wellbeing, and pregnancy and birth outcomes in various settings.

**Method:** A systematic search of PubMed, EMBASE Classic + EMBASE (Ovid), MEDLINE In-Process and Non-Indexed Citations, MEDLINE Daily, and MEDLINE (Ovid), Cumulative Index to Nursing & Allied Health Plus (CINAHL via EBSCO) and Cochrane library databases was conducted to identify studies on stress reduction relaxation interventions in pregnant women. The outcomes of interest were maternal mental health (stress, anxiety, and depression), pregnancy outcomes (gestational age, labor duration and mode of delivery) and birth outcomes (birth weight, APGAR score and term or preterm delivery). Randomized controlled trials or quasi-experimental studies with stress reduction relaxation interventions during pregnancy and ever published in English globally were eligible for inclusion. Studies with interventions in high-risk pregnancies, those including psychotropic medications, or interventions at the onset of labor and delivery were excluded. All studies were screened for quality and risk of bias. We conducted meta-analyses, using random-effects models, for three outcomes for which there was sufficient information: maternal depressive symptoms, perceived maternal stress; and birth weight.

**Result:** Nineteen studies were eligible for analysis. The studies sampled 2395 pregnant women, mostly aged between 18 and 39 years. The interventions applied were yoga therapy, music therapy, progressive muscular relaxation (PMR)/guided imagery/deep breathing exercises, mindfulness or hypnosis. The meta-analyses showed that the interventions were effective in improving maternal depressive symptoms (−2.5 points, [95% confidence interval (CI) -3.6, -1.3]) and stress symptoms (−4.1 points, [95% CI -8.1, -0.1]) during pregnancy. There was no effect of the interventions overall on birth weight (45 g, 95% CI -56, 146); however, PMR in two studies increased birth weight (181 g, 95% CI 25, 338) whereas music therapy and yoga had no benefit. Narrative syntheses of outcomes that were not amenable to meta-analysis indicated beneficial effects of music interventions on APGAR score (n=4 studies) and gestational age at birth (n=2 studies). Interventions were also reported to significantly increase spontaneous mode of delivery (n=3 studies) and decrease the rate of instrumental virginal delivery by 5%, caesarean section by 20% and duration of labor (n=2 study).

**Discussion:** Adverse life experience during pregnancy impairs the normal adaptive changes supposed to maintain normal homeostasis during pregnancy and results in increased risk of stress, anxiety and depression. This imbalance results in increased stress hormone in the maternal-fetal circulation which is harmful to the mother and her fetus leading for adverse pregnancy and birth outcomes. Stress reduction relaxation intervention restores the normal homeostasis in pregnancy and improves normal biological and psychological wellbeing and consequently improves pregnancy and birth outcomes.

**Conclusion:** In addition to benefits for mothers, relaxation interventions hold some promise for improving newborn outcomes; therefore, this approach strongly merits further research.

## Introduction

Mental wellbeing is a state whereby a person realizes his/her own abilities, can function productively, and can cope with normal daily stressors [1]. Maternal stress and disturbances in mental wellbeing are common problems during pregnancy and after childbirth [2]. The mental wellbeing of pregnant women can be challenged by the physiological, psychological and social and role changes during pregnancy, and may be aggravated or mitigated by the level of personal and social support [3]. Risk factors for symptoms of stress during pregnancy include poor social and financial support, fear/worry about pregnancy and birth outcomes, previous bad obstetric experiences, physiological changes of pregnancy, and the change in role associated with motherhood [2–4]. Empirical evidence suggest that 15–25% of women experience high levels of anxiety or depressive symptoms during pregnancy [2,3]. The estimate for the magnitude of prenatal depression and anxiety in low- and middle-income countries also falls within the same range [5–7].

Maternal stress during pregnancy increases adverse pregnancy and birth outcomes [3,8], such as maternal poor mental health, premature delivery, low birth weight (LBW), as well as increasing the risk of maternal and neonatal mortality [3,9]. Many risks for under-five malnutrition originate in prenatal life, and LBW affects approximately 15% of infants, mostly in low- and middle-income countries (LMICs) [9,10]. Early life malnutrition is a risk for later life abdominal adiposity, obesity and non-communicable disease (NCD) in countries undergoing nutrition transition [9]. The effect of psychological stress during pregnancy on neonatal birth outcomes was reported among women of different ethnic, socio-economic and cultural backgrounds [11– 13]. Prenatal maternal stress symptoms adversely influence not only fetal growth and development but also the physical and mental health, and neuro-behavioral development of the offspring [14].

Various interventions, including psychotropic medication, counseling and psychosocial support, as well as relaxation therapies, have been tested to improve the psychological wellbeing of pregnant women. However, no comprehensive review was available to show the synthesized evidence on the effectiveness of relaxation interventions on maternal and neonatal outcomes. This paper therefore aimed to synthesize evidence on the effects of relaxation interventions on stress during pregnancy (self-report, physiological or biochemical) and maternal and neonatal outcomes.

## Methods

### Protocol and registration

The protocol for this review was registered at PROSPERO International prospective register of systematic reviews and can be accessed at: https://www.crd.york.ac.uk/prospero/display_record.php?ID=CRD42020187443

### Article Selection

The review process followed the Preferred Reporting Items for Systematic Review and Meta-Analysis (PRISMA) guideline [15]. To identify relevant articles, a three-step search strategy was employed. In the first step, key free text and MeSH terms were identified and developed, and then a comprehensive search was conducted using the following five databases: PubMed, EMBASE Classic + EMBASE (Ovid), MEDLINE in-process and non-indexed citations, MEDLINE daily, and MEDLINE (Ovid), Cumulative Index to Nursing & Allied Health Plus (CINAHL via EBSCO), Cochrane library. In addition, a manual search was conducted to identify further additional relevant studies from the reference lists of identified studies. Unpublished and grey literatures were excluded.

The search terms were developed based on the population (“Pregnant women” OR “pregnancy” OR “prenatal” OR “prenatal care” OR “mother” OR “antenatal” OR “antenatal care” OR “maternal” OR “maternal care”) AND type of intervention (“Relaxation therapy” OR “Mindfulness therapy” OR “Progressive muscle relaxation (PMR) therapy” OR “Music therapy” OR “Exercise therapy” OR “deep breathing relaxation therapy” OR “Meditation therapy” OR “hypnosis therapy” OR “relaxation lighting”), AND Outcome of interest (“Stress” OR “distress” OR “anxiety” OR “depression” OR “Birth-weight” OR “birth weight” OR “birth outcome” OR “Apgar”, “Apgar score”, “Gestation”, OR “Gestational age at birth”). Only Randomized Controlled Trials (RCT) and quasi-experimental studies were included; case reports, cross-sectional studies, and editorials or opinion pieces were excluded.

Studies were eligible if they employed randomized controlled trial (RCT) or quasi-experimental designs, were published in English, intervened in apparently healthy pregnant women, and reported any of the outcomes of interest specified in our search strategy. Case reports, cross-sectional studies, and editorials or opinion pieces were excluded. Studies that involved women at risk for preterm delivery, complicated or high-risk pregnancies, or women taking psychotropic or other medications, or where interventions were provided only at the onset of labor or after delivery, were also excluded.

### PICO

#### Population

apparently healthy pregnant women.

#### Intervention/exposure

stress reduction and/or relaxation therapy. Any form of relaxation intervention, whether mind-based (tapes, music) or physical/body-based (massage, stretch, or exercise) including Progressive Muscle Relaxation (PMR) and deep breathing exercises, that were applied with the aim of promoting relaxation and reducing stress.

#### Comparators/controls

pregnant women who did not receive a stress-reduction intervention or who received treatment as usual.

#### Outcomes

the main outcomes were measures of stress/relaxation (self-report, physiological, or biochemical) or mental health or depressive or anxiety symptoms or pregnancy outcomes (birth weight, Apgar score, gestational age at birth, mode of delivery, and duration of labor).

#### Timing of outcome measures

studies that measured the outcome during, immediately after, or some weeks or months after the intervention was delivered were included.

### Study screening process

The literature search was concluded on 6 July, 2022. To decide on inclusion, the studies were first screened by titles and then by abstracts using the eligibility criteria. Secondly, full texts of the selected studies were assessed based on the inclusion and exclusion criteria. Two authors (MA and BD) screened all the articles for eligibility. Any queries were discussed with one additional author (AW) to reach consensus. The screening process and reasons for exclusion of studies were documented.

### Methodological quality assessment

Two independent assessors (BD and MA) evaluated the methodological quality of studies in terms of randomization, masking, and availability of descriptions for withdrawal and drop out of all participants based on the modified Jadad scoring scale [16] and the modified Delphi List Criteria [17] to assess the overall quality of the studies. Using the Cochrane collaboration’s assessment checklist [18], risk of bias was assessed and rated as low, high or unclear for individual elements relating to five domains (selection, performance, attrition, reporting, and other). The criterion on blinding was excluded as it is usually impossible to conduct relaxation therapy while blinding the participant or the care providers.

### Data extraction

The findings were extracted using a standard data abstraction form. Data were extracted in two phases. In the first phase, citation details (author name, publication year, design, sample size, setting, population, intervention, comparison, and outcomes) were extracted. In the second phase, the intervention results by group were extracted.

### Strategy for data synthesis

To obtain the pooled effects of the intervention, we conducted meta-analyses on the following outcomes for which there was adequate information: maternal depressive symptoms, maternal stress symptoms and birth weight. We used the mean difference (MD) with the reported Standard Deviation (SD) of the outcomes as a measure of effect size for each of the included studies. For the meta-analysis, the raw mean difference (D), with 95% CI across studies that measured the same outcome (depression with Edinburgh Postnatal Depression Scale (EPDS) or stress with Perceived Stress Scale (PSS), and birth weight in grams) was examined and presented. Sub-group analyses were performed to examine the existence of significant differences among studies that used different relaxation interventions for any given outcome of interest. We assessed heterogeneity with the Cochrane’s Q test and tau-squared (T^2^) and measured inconsistency (the percentage of total variation across studies due to heterogeneity) of effects across relaxation interventions using the I^2^ statistic. Publication bias was assessed using regression based on Egger’s test. For all meta-analyses, random effects models using restricted maximum likelihood estimates (REML) were employed. Statistical significance was defined as *P* < 0.05. Stata version 16 software was used for the meta-analyses.

For outcomes where a meta-analysis was not possible because of inadequate number of studies and small sample size, a narrative synthesis of the reviewed articles on the effect of the interventions on each outcome of interest was performed and reported.

## Results

### Search result: Final reviewed studies

A total of 19 studies were included in the systematic review (**Error! Reference source not found**.).

One of the studies was a non-randomized controlled trial while the other 18 were RCTs on the effect of relaxation interventions during pregnancy on depressive symptoms, stress or anxiety symptoms in pregnant women, as well as on birth outcomes including birth weight, Apgar score, and gestational age at birth. From the 19 trials, 12 trials that reported similar measurements and outcomes, and provided sufficient information were included in the quantitative meta-analysis. Four of those trials assessed maternal depression using EPDS, six trials examined maternal stress among the mothers using PSS-10 and five of the trials examined birth weight. Four trials reported on Apgar score, two trials reported on gestational age, three trials on mode of delivery, and one trial on duration of labor; these were not included in the meta-analyses as they did not provide sufficient information or did not use similar measurements.

### Study contexts/settings

Three studies were from India, a lower middle-income country, and the rest were from upper-middle- (MIC) or high-income (HIC) countries. Reviewed trials on the effect of relaxation interventions on maternal stress, anxiety or depression included one study each from the United States, UK, Switzerland, Greece, Malaysia, Iran, Turkey, and China, two studies from India, and five studies from Taiwan. Studies on relaxation therapy and birth outcomes included one trial each from India, Iran, Thailand, and Spain. There were no published studies from Africa or from other low-income countries (LIC) in other global regions.

### Risk of bias within and across studies

Most studies had low risk of selection bias, random allocation and concealment bias, and other sources of bias. Most studies had unclear risks of bias for reporting bias (selective reporting of outcomes). One study had high risk of bias as there was no random generation and assignment, as well as incomplete reporting of outcome data [19]. Supplementary Table 1 shows the risk of bias assessment findings for each of the studies.

### Types of relaxation intervention

The reviewed articles used one or a combination of the following relaxation therapies: yoga therapy, music therapy, progressive muscular relaxation (PMR)/guided imagery/deep breathing exercises, mindfulness or hypnosis. Four of the RCTs applied yoga as a relaxation intervention. The yoga interventions included a 6 – 8 -week course of a combination of prenatal yoga techniques specifically adapted to pregnant women covering relaxed breathing, body awareness, and non-strenuous postures for different stages of labor. Music therapy was applied in seven of the RCTs. Five RCTs used PMR, Guided imagery (GI) or deep breathing exercise. Two of the RCTs used mindfulness therapy, and one study used hypnosis as a relaxation intervention. Table 1 contains more information on the included studies.

**Table 1:** Description and summary of the results of the studies included in the systematic review and meta-analysis.

**Table 2:**
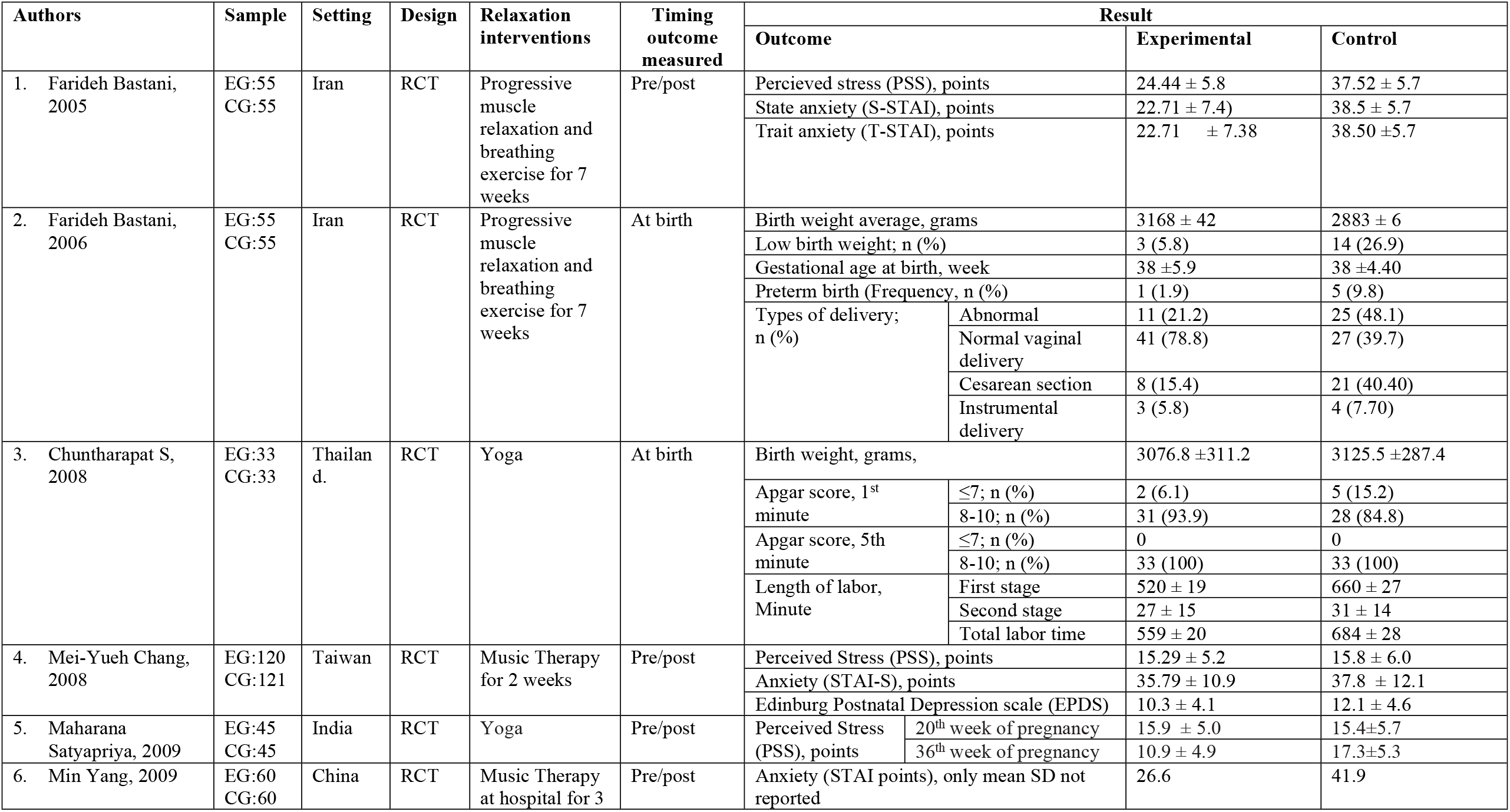
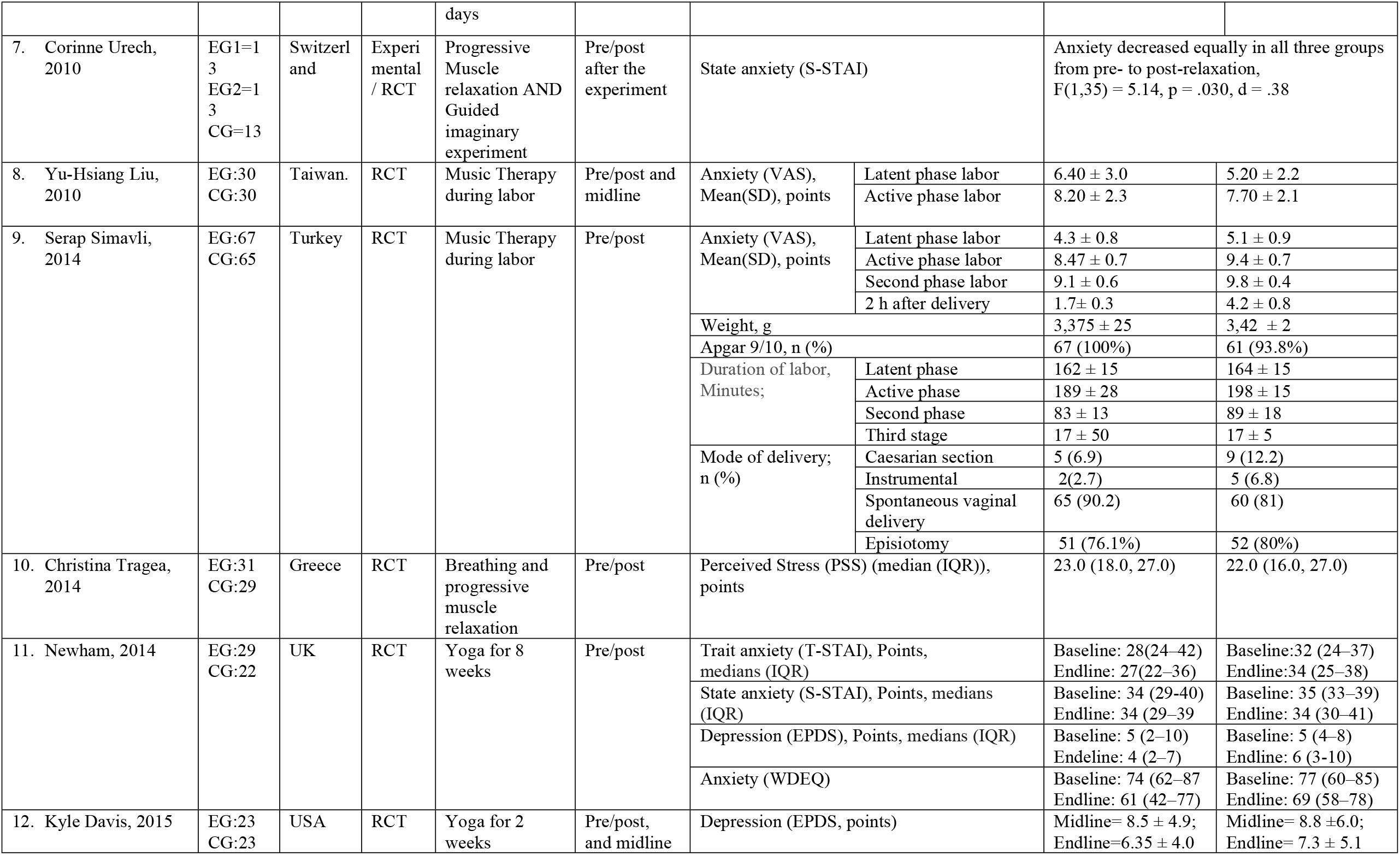
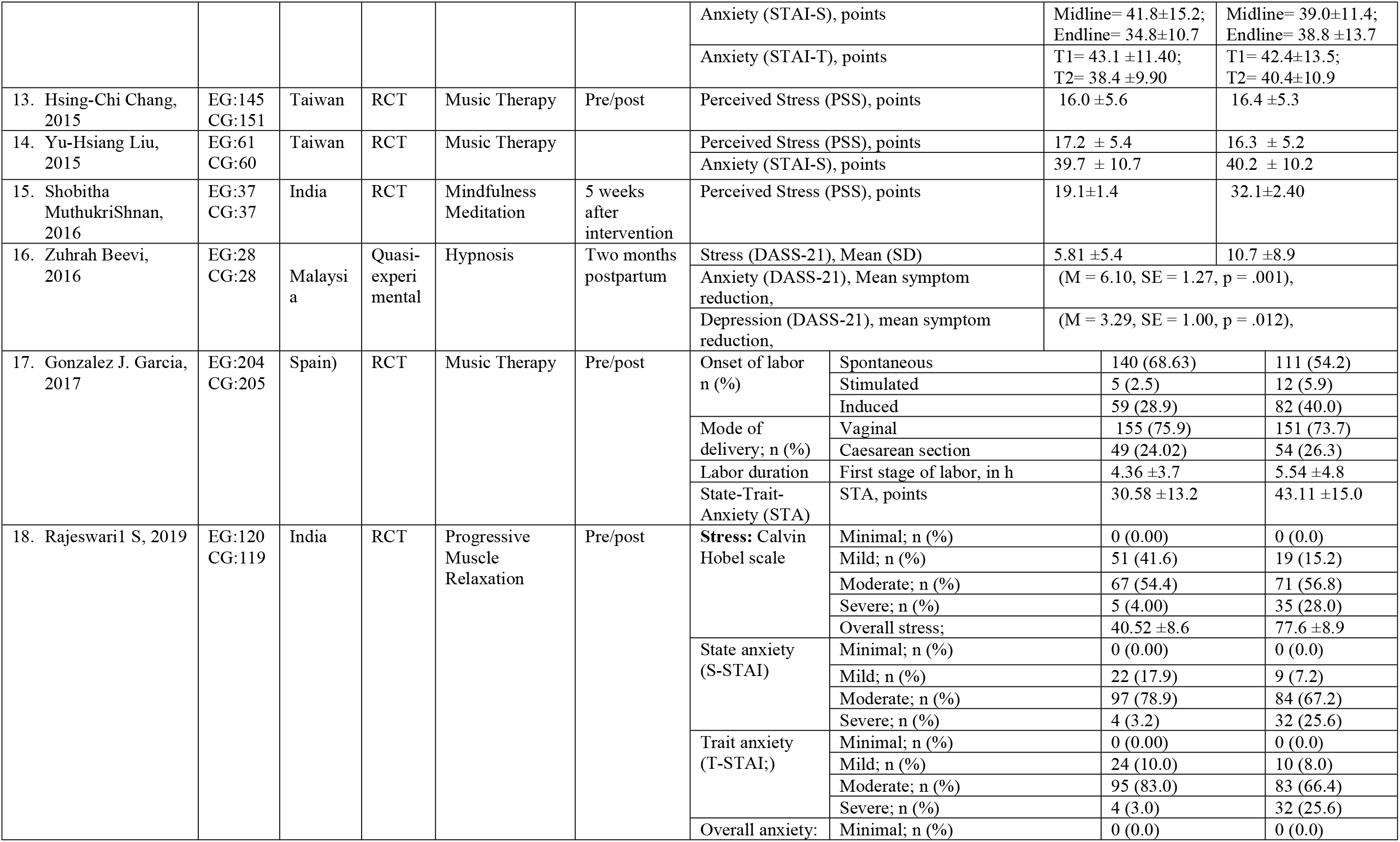
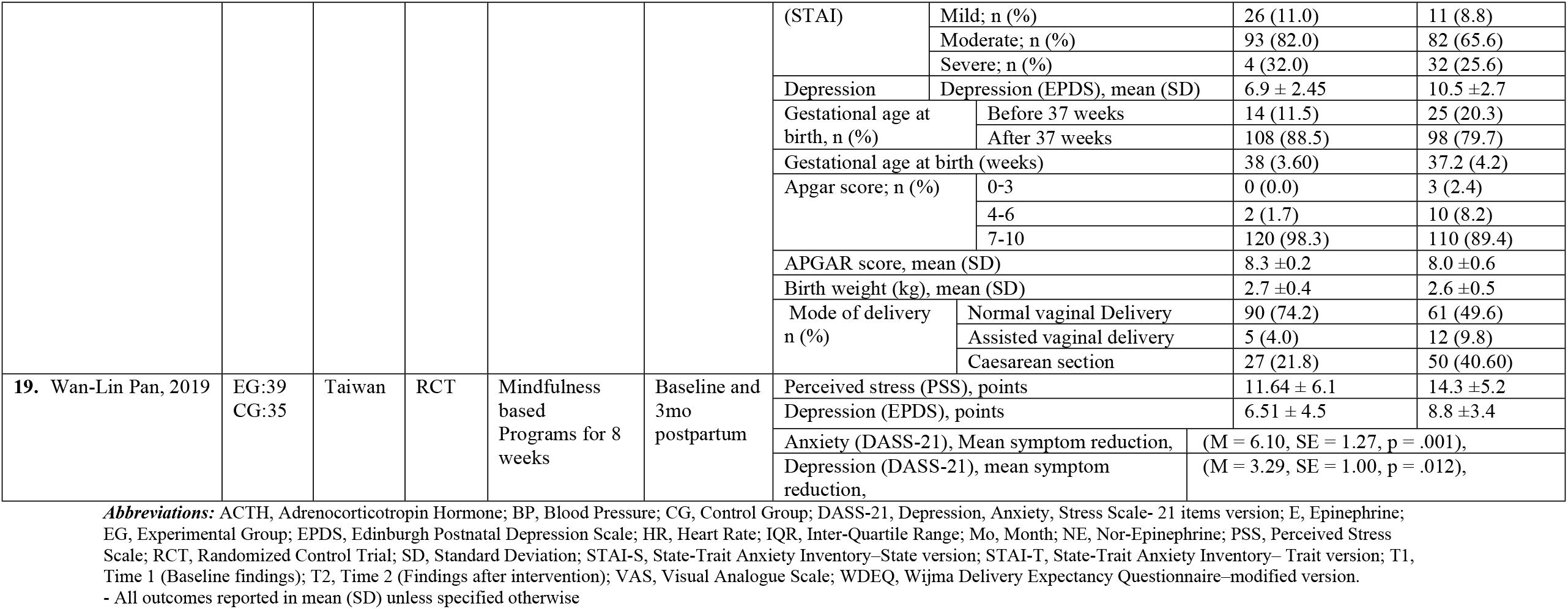
Description and summary of the results of the studies included in the systematic review and meta-analysis.

### Intervention outcomes

#### Maternal mental health symptoms during pregnancy

Outcomes for the effect of relaxation intervention during pregnancy on maternal mental health were stress, anxiety and depressive symptom scores. Different scales were used to measure the levels of maternal stress, anxiety, and depressive symptoms and the findings are summarized below ***(Table 1)***.

#### Maternal depressive symptoms

Four trials examined the effectiveness of relaxation interventions on maternal depressive symptoms measured using the Edinburgh Postnatal Depression Scale (EPDS) [20–23]. An RCT in Taiwan allocated pregnant women into an intervention group that received a Mindfulness-Based Childbirth and Parenting (MBCP) intervention for 8 weeks with routine clinical care or a control group given routine care only. Another RCT provided a group of mothers with a music CD to listen to daily for two weeks, while control mothers were provided with routine prenatal care [21]. In an RCT in India, PMR was provided for randomly selected primigravidae mothers that were divided into experimental and control groups [23]. An RCT in the United States randomized women to receive Yoga compared to women that received routine antenatal care [20].

A meta-analysis of the four RCTs was performed. The relaxation interventions resulted in a significant reduction in mean EPDS scores (mean difference *=* -2.47, 95% CI [-3.64, -1.31], p<0.001) between the mothers who received the interventions and those who did not **(Error! Reference source not found.)**. The studies were highly heterogeneous (Q=10.57, I^2^=67.2%, p=0.01). Publication bias of small study was not significant (beta1=2.43, p=0.09).

#### Maternal stress symptoms

Nine RCTs reported the effectiveness of relaxation in alleviating maternal stress symptoms. Three RCTs in Taiwan reported a significant improvement in stress in mothers who received music therapy compared to the controls [21,24,25]. In India, lower mean perceived stress score was reported among mothers who received mindfulness-based meditation compared to those who received routine antenatal care [26]. Another RCT conducted in Taiwan reported similar findings in which mindfulness-based childbirth and parenting program resulted in a significant reduction in maternal stress [22]. Integrated yoga practice and guided yogic relaxation were examined for their effectiveness in alleviating maternal stress in India [29].

The perceived stress scale was also used in an RCT conducted in Greece to examine the effect of PMR/ deep breathing exercise between experimental (n=31) and control (n=29) groups with measurements taken before and after intervention. There was a significant difference in post-intervention stress between the groups. Additionally, the mean pre-post difference was higher in the intervention group (−3.68 ± 1.80) than the control group (−0.45±1.80) [27]. The effectiveness of PMR/ deep breathing exercise in reducing maternal stress was also reported in two other studies conducted in Iran [28] and India [23]. One non-randomized trial that examined the effectiveness of hypnosis on maternal stress reported that the intervention group (n=28) had less stress than the control group (n=28) (5.8 ±5.4 vs 10.7±9) [19].

A meta-analysis of six RCTs that examined the effect of relaxation interventions on perceived stress showed that the relaxation interventions resulted in a significant reduction in stress among pregnant mothers (mean difference = -4.09 95%CI [-8.10, -0.07], p<0.001). There was a high level of heterogeneity among the studies (I^2^=98.02 %). Subgroup analysis by the type of relaxation interventions revealed a significant difference among the different interventions (Q=434.20, p<0.001); meditation caused the highest reduction in maternal PSS scores while music therapy resulted in the lowest reduction in the PSS scores **(Error! Reference source not found.)**.

#### Maternal anxiety symptoms

A narrative synthesis of the effect of relaxation on maternal anxiety indicated inconsistent findings. Studies in Iran and India, reported significant reductions in maternal anxiety symptoms in the experimental groups that received PMR compared to the control groups [23,28].

Conversely, two RCTs conducted in USA and UK found no significant difference between groups that received yoga compared to control groups regarding their anxiety levels [20,30]. However, the UK study reported a significant reduction in women’s anxiety toward childbirth measured using the Wijma Delivery Expectancy Questionnaire (W-DEQ) in the mothers who received yoga [30].

Anxiety levels were reported to be significantly lower in groups that received music therapy compared to controls in three separate studies conducted in Taiwan and China [21,25,31]. The effect of music therapy on maternal anxiety measured by visual analogue scale was also reported in two studies. Both studies reported a significant reduction in maternal anxiety in the intervention group compared to the control group [32,33]. A study in Turkey reported a reduction in anxiety symptoms in all phases of labor, while another study conducted in Taiwan reported a difference during the latent phase of labor, but not during the active phase of labor [32,33]. Meta-analysis was not performed for maternal anxiety as there were an inadequate number of studies that reported the outcome with comparable measures.

### Newborn birth outcomes

Five RCTs investigated the effect of relaxation interventions on newborn outcomes [23,32,34– 36]. All five trials examined the effects of the intervention on birth weight.

Two studies conducted in India indicated that PMR/deep breathing exercise had a significant effect on the birth weight of the infants and decreased rates of LBW [23,34]. On the other hand, studies conducted in Turkey and Spain for music therapy and in India for yoga reported the interventions had no significant effect on birth weight [32,35,36].

Our meta-analysis of five RCTs on the effect of the interventions on birth weight showed that the relaxation interventions resulted in no significant differences (*mean difference* = 45.2, 95% CI [-55.6, 146.0] g, p=0.38) however there was a significant heterogeneity among the studies (I^2^=73.54%) (**Error! Reference source not found.)**. Subgroup analysis revealed there was a significant group difference among the studies that used different interventions (Q=5.98, p=0.05). PMR interventions increased birth weight, whereas music and yoga had no significant effects.

APGAR score was measured in four trials. A study in Turkey reported that 100% of neonates born to mothers who received music therapy had Apgar scores of 9/10 while only 93.8% of neonates born to mothers in the control group scored 9/10 (p=0.05) [32]. Another study in Spain reported no statistically significant difference in the Apgar score of the neonates between the music therapy group and the control group [36]. Additionally, neither yoga nor PMR significantly affected Apgar score [23,35].

An RCT in Iran examined the effects of PMR on gestational age at birth and the rate of preterm birth (experimental and control groups n=55 each). The authors reported no significant difference in gestational age at birth between the two groups post-intervention [34]. In contrast, another RCT in India reported a significant effect of PMR on gestational age at birth and rate of preterm birth [23]. The authors reported that mean (SD) gestational age at birth for mothers in the experimental group was significantly greater than in the control group (38.0 (3.6) weeks vs 37.2 (4.2) weeks, t=1.97, *p*=0.04). The same RCT also reported that the percentage of births prior to 37 weeks of gestation was significantly lower in the experimental (11.5%) compared to the control group (20.3%) (F2=6.08, *p*=0.014) [23].

### Obstetric outcomes

Four of the RCTs assessed the effects of relaxation on obstetric outcomes. In Turkey, it was reported that music therapy had no effect on the mode of delivery [32]. In contrast, Iranian mothers who received PMR had fewer instrumental deliveries and caesarean sections than mothers in the control group (21.2 % vs 48.1%, P=0.002) and an increased rate of spontaneous vaginal delivery (78.8% vs 39.7, P=0.01) [34]. Similarly, there was a significant reduction in assisted deliveries and caesarian deliveries in mothers who received PMR in India [23]. The same trial reported there a statistically significant difference in the rate of induced labor among mothers in the PMR group and the control group (F2=5.50, p=0.019) [23].

One study examined the effect of music therapy on duration of labor among mothers in Turkey. They reported that the experimental group had significantly shorter mean (SD) labor time for active first stage 189 (28) minutes vs 198 (15) minutes and second stages 83 (13) minutes vs 89 (18) minutes of labor than the control group [32]. An RCT in Thailand also reported a shorter first stage labor 520 (186) minutes Vs 660 (273) minutes and total duration of labor 559 (203) minutes vs 684 (276)minutes in the yoga group compared to the control group [35].

## Discussion

This paper reviewed available evidence on the effect of various relaxation interventions on maternal stress, mental health, and obstetric and newborn outcomes. The relaxation interventions included were music therapy, PMR, yoga, deep breathing exercise, meditation, and mindfulness interventions, and the trials examined the effects on reducing stress during pregnancy, and improving maternal wellbeing, obstetric and birth outcomes.

The findings of this review provide a better understanding on the effect of different types of relaxation interventions in improving maternal stress, anxiety and depression. The meta-analysis showed that relaxation interventions in general are effective in reducing perceived stress and depressive symptoms during pregnancy. MBCP, Music therapy, and PMR were shown to be effective in reducing maternal depressive symptoms [21–23] whereas music therapy [21,24,25], meditation [26], mindfulness-based childbirth and parenting [22], PMR [23,27,34], yoga [29], and hypnosis [19] improved stress symptoms during pregnancy. Maternal anxiety symptoms were found to be lower with the application of PMR [23,28] and music therapies [21,25,31–33].

In normal circumstances, pregnant women undergo some biochemical and physiological adjustments in the endocrine, nervous and immune systems, all of which contribute to the development of normal pregnancy and birth outcomes [37,38]. To achieve these adjustments, maternal stress responses and inflammatory responses are suppressed during pregnancy because of a down-regulated hypothalamic-pituitary-adrenal (HPA) axis and the shift of the immune system to favor anti-inflammatory responses [39]. However stress due to various psycho-social stressors during pregnancy affects these normal biochemical and physiological adaptations which are supposed to support optimal maternal health and wellbeing, and the offspring’s prenatal and postnatal growth and development [37–39]. Thus, through its counter-effect on maternal stress response, relaxation interventions help pregnant women maintain this normal homeostasis/adjustment. Relaxation improves breathing and helps the body to release muscle tension and to regain calmness and a sense of rest with improved heart rate, blood pressure, digestion, absorption, and blood circulation [40–42]. Through these mechanisms, relaxation helps to reverse the development or worsening of symptoms of anxiety and depression in pregnant women.

The results from the meta-analysis showed that relaxation interventions have no overall effect on birth weight. However, there were differences in the outcome depending on the type of intervention: PMR had positive effects on birth-weight [23,34] while music and yoga had no effect [32,35,36]. Studies on the other outcomes including gestational age, mode of delivery, and duration of labor resulted in improved gestational age and spontaneous delivery and reduced instrumental or cesarean delivery. Though the findings on obstetric and birth outcomes are inconclusive at the moment, the interventions showed promising effects in improving obstetric and birth outcomes. The current trials included in the review on obstetric and birth outcomes had small sample sizes and are heterogeneous which could partly explain the lack of significant effects on obstetric outcomes. Thus, there is a need for further evidence on the effect of relaxation interventions on obstetric and birth outcomes to get adequate power so as to reach robust conclusions.

There is already a body of evidence that stress, anxiety, and depression, through their direct or indirect effects, result in adverse neonatal and obstetric outcomes [8,43,44]. Through its indirect effect, stress during pregnancy reduces maternal capacity for self-care during pregnancy, and may result in sub-optimal preparation for delivery and infant care during the post-natal period [45]. Moreover poor self-care during pregnancy, sleep disturbance, increased risk for maladaptive behaviors, poor help-seeking behavior and poor nutrition as a result of poor maternal wellbeing contributes to adverse pregnancy and birth outcomes [8,46,47]. These adverse effects of maternal stress during pregnancy may be transmitted to the offspring even if fetal growth and gestation are not affected, as demonstrated by research on epigenetic effects [14]. Moreover stress hormone-related suppression of the immune system contributes to increased risk of maternal infection/illness which may further complicate pregnancy and birth outcomes [46,48].

As a direct effect, stress during pregnancy activates the stress response mechanism through the activation of the HPA axis and subsequent release of corticotrophin releasing hormone (CRH) leading to increased release of cortisol to the fetal circulation which will result in decreased supply of oxygen and essential nutrients from the mother to fetus [46,49,50]. This may contribute to preterm birth, low birth-weight and other poor obstetric outcomes [51–54]. Fetal exposure to glucocorticoids during pregnancy programs the development of the fetal stress response and can thus affect the health, growth and development of the offspring across the life span [55]. In summary, the up-regulated HPA axis and down-regulated immune system with an increased inflammatory response during pregnancy may all affect fetal and postnatal growth and development of the offspring.

Thus the mechanisms through which relaxation interventions could improve pregnancy and birth outcomes involve an interplay between psychological, biochemical, physical, and endocrine effects [56–60]. In previous studies, relaxation interventions have shown reduced release of these stress hormones (cortisol, epinephrine, norepinephrine, and adrenocorticotrophic) thereby preventing the activation of the HPA axis and its effect on the placental circulation, a key component of maternal-fetal communication [58–60]. To see stronger and robust findings on the effect of relaxation interventions on obstetric and birth outcomes there is a need for additional primary studies with adequate power and sample size to contribute to the current gab in evidence to make a conclusive decision.

## Limitations

As strength of this work, we have included studies form different forms of relaxation intervention and have done both narrative as well as pooled meta-analysis based on data availability. The interpretation of the findings of the systematic review and meta-analysis should be taken with some careful considerations. The first is the limited number of studies conducted on the topic and the small number of studies providing evidence for the effectiveness of some of the interventions. Data on the effect of the relaxation interventions on neonatal outcomes other than birth weight was very limited and was insufficient to conduct a meta-analysis. In addition, the review team did not contact authors when there was missed information on the reported studies. This may have impacted the amount of information from the studies available for narrative synthesis as well as meta-analysis.

Most of the studies included in this review are from middle- or high-income countries with the exception of India, which is a lower middle-income country. This indicates that evidence is still lacking for the effectiveness of these interventions in low-income countries, including those in sub-Saharan Africa. It is crucial to establish an evidence base in sub-Saharan African countries as populations in these countries experience persistent high levels of fetal and child undernutrition which continue to impair health and development of millions of children.

### Conclusion and Recommendation

The results of the review in general indicate that, in addition to benefits for mothers, relaxation interventions hold some promise for improving newborn outcomes; therefore, this approach strongly merits further research. Considering the high magnitude of perinatal mental health and psychological problems, the high burden of obstetric complications and the associated maternal and neonatal morbidity and mortality, the results of this review indicate sets of complementary treatment mechanisms for tackling the problems and contributing to the achievement of the sustainable development goals, specifically the third goal of achieving good health and wellbeing. Their relative cost-effectiveness, ease, and relative absence of adverse and teratogenic effects in comparison to pharmacological treatments favor the application of one or combination of the relaxation interventions in this population group. Relaxation interventions are low-intensity and may be more scalable than individualized psychological interventions in resource-limited settings.

Therefore, we recommend that these relaxation interventions be evaluated for their effectiveness in LMICs so that relaxation can be integrated in the routine clinical care of pregnant women. Further evaluating the interventions in these settings would also be beneficial to understand whether, in places with massive food insecurity and high burden of infections, which affect both maternal and infant health, a relaxation intervention could mitigate the effect of these stressors. We recommend routine consideration of the mental health of pregnant women during antenatal care visits. This recommendation is in line with the global initiatives such as the World Health Organization’s mental health Gap Action Program which are based on integration of mental health care into all health settings, including maternal care. In addition, we recommend further studies to be conducted on the effectiveness of these interventions on maternal, obstetric, and neonatal outcomes in low-income countries including in sub-Saharan Africa.

**Figure 1:**
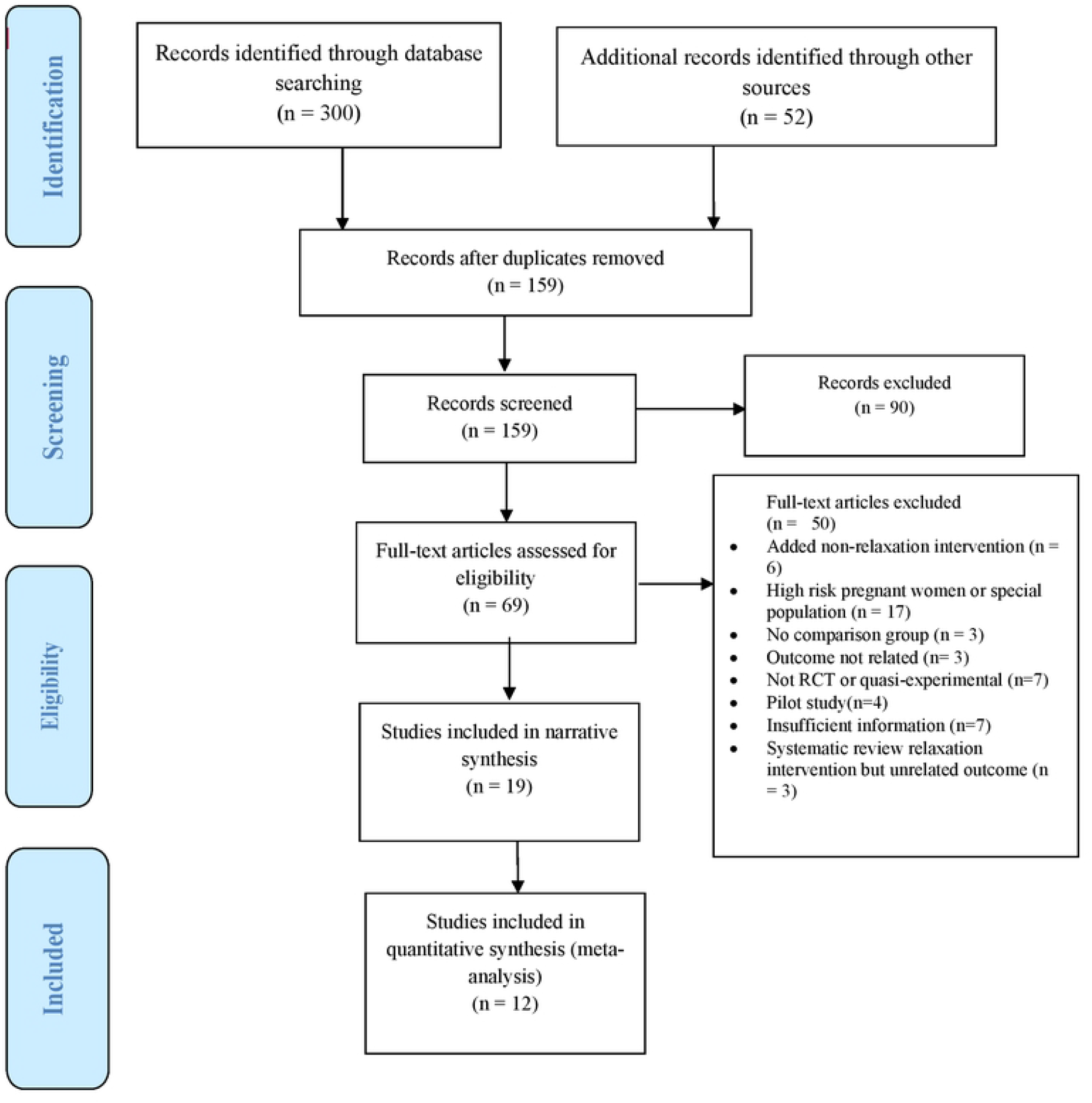
PRISMA flow chart showing literature search results and study selection process

**Figure 2:**
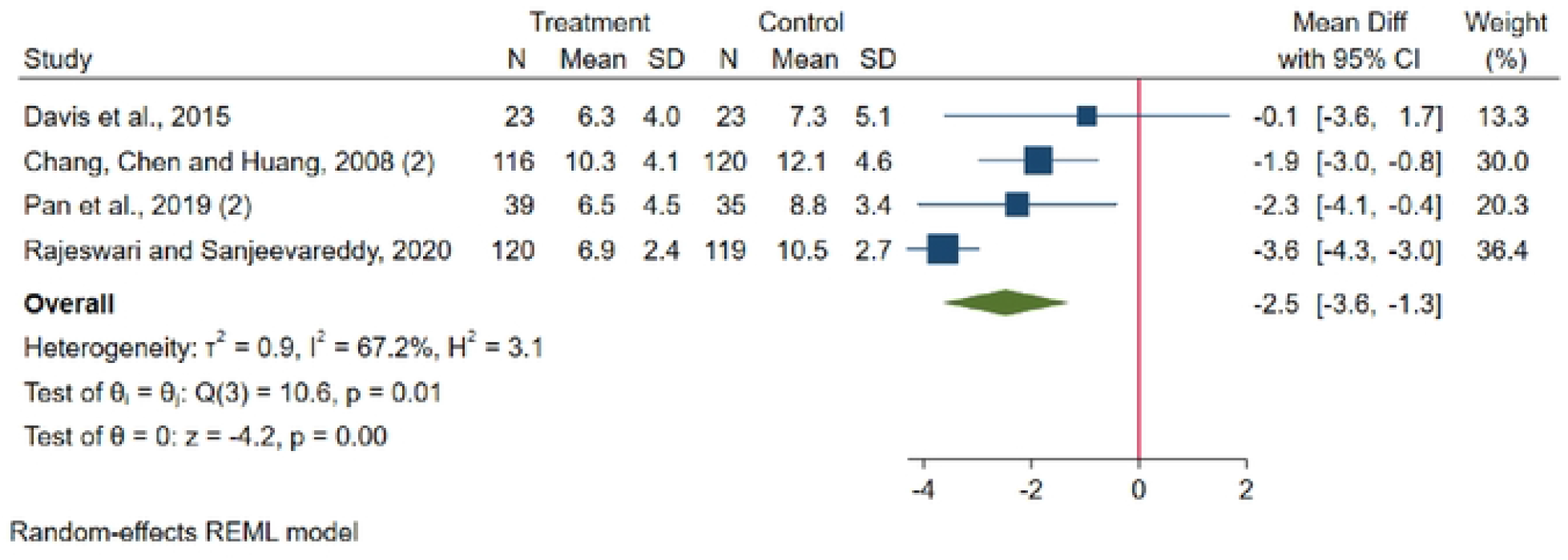
Forest plot for the raw mean difference in Edinburgh postnatal depression scale score between the intervention and control groups.

**Figure 3:**
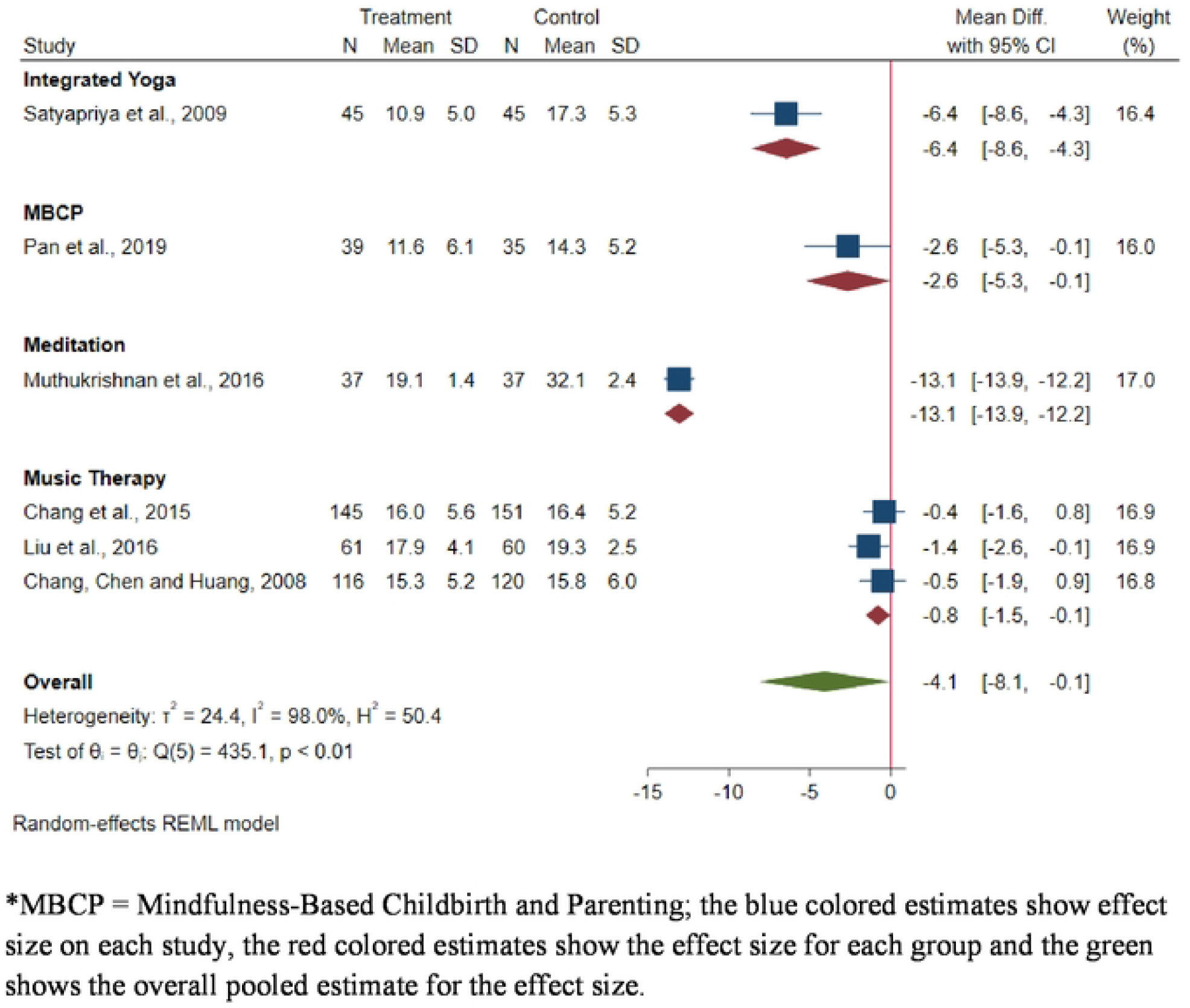
Forest plot and subgroup analysis for raw mean difference of studies for the effect of relaxation interventions on maternal stress measured using perceived stress scale.

**Figure 4:**
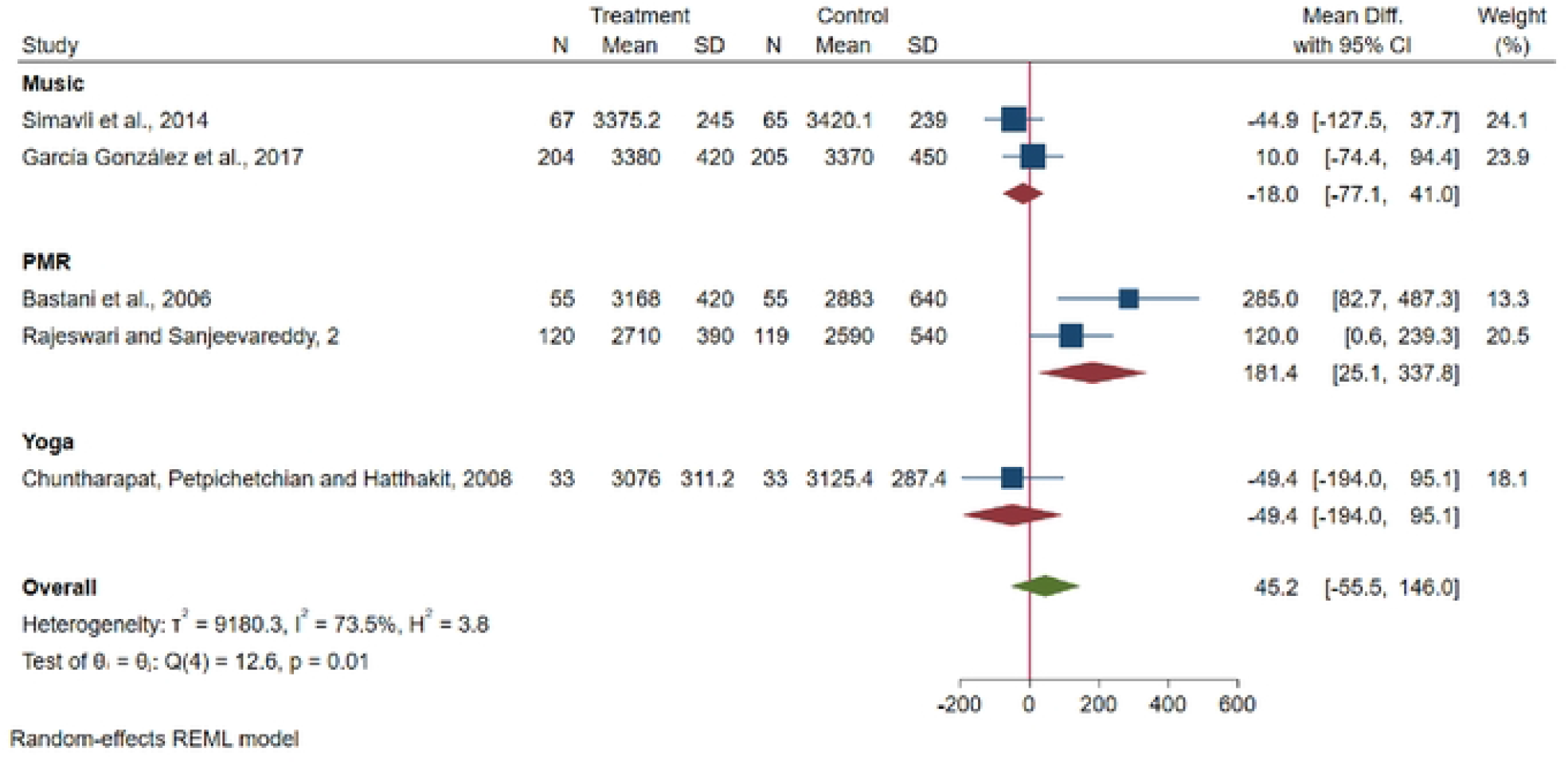
Forest plot and subgroup analysis for raw mean difference of studies for the effect of relaxation interventions on birthweight (g).

## Data Availability

All relevant data are within the manuscript and its Supporting Information files.

## Notes

### Competing Interest Statement

The authors have declared no competing interest.

### Funding Statement

The author(s) received no specific funding for this work

### Author Declarations

This study does not require IRB as it is a secondary data analysis

## References

1. World Health Organization. Promoting Mental Health: Concepts, practice, summary report. 2004.

2. Figueiredo B, Conde A. Anxiety and depression in women and men from early pregnancy to 3-months postpartum. 2011;247–55.

3. Entringer S, Buss C, Wadhwa PD. Psychoneuroendocrinology Prenatal stress, development, health and disease risk : A psychobiological perspective — 2015 Curt Richter Award Paper. Psychoneuroendocrinology. 2015;62:366–75.

4. Lee AM, Lam SK, Marie S, Mun S. Prevalence, Course, and Risk Factors for Antenatal Anxiety and Depression. 2007;110(5):8–10.

5. Tesfaye Y, Agenagnew L. Antenatal Depression and Associated Factors among Pregnant Women Attending Antenatal Care Service in Kochi Health Center, Jimma Town, Ethiopia. J Pregnancy. 2021;2021.

6. Lodebo M, Birhanu D, Abdu S, Yohannes T. Magnitude of Antenatal Depression and Associated Factors among Pregnant Women in West Badewacho Woreda, Hadiyya Zone, South Ethiopia: Community Based Cross Sectional Study. Depress Res Treat. 2020;2020.

7. Fisher J, de Mello MC, Patel V, Rahman A, Tran T, Holton S, et al. Prevalence and determinants of common perinatal mental disorders in women in low-and lower-middle-income countries: A systematic review. Bull World Health Organ. 2012;90(2):139–49.

8. Dadi AF, Miller ER, Bisetegn TA, Mwanri L. Global burden of antenatal depression and its association with adverse birth outcomes: An umbrella review. BMC Public Health. 2020 Feb 4;20(1).

9. Wells JC, Sawaya AL, Wibaek R, Mwangome M, Poullas MS, Yajnik CS, et al. The double burden of malnutrition: aetiological pathways and consequences for health. Lancet. 2020;395(10217):75–88.

10. World Health Organization. Global nutrition targets 2025: low birth weight policy brief. World Health Organization. 2014.

11. Schetter CD. Psychological Science on Pregnancy : Stress Processes, Biopsychosocial Models, and Emerging Research Issues. 2011;

12. Guardino CM, Dunkel Schetter C, Bower JE, Lu MC, Smalley SL. Randomised controlled pilot trial of mindfulness training for stress reduction during pregnancy. Psychol Heal. 2014;29(3):334–49.

13. Hobel C, Culhane J. Life and Fitness Role of Psychosocial and Nutritional Stress on Poor Pregnancy Outcome 1. 2018;(May):1709–17.

14. Van den Bergh BRH, van den Heuvel MI, Lahti M, Braeken M, de Rooij SR, Entringer S, et al. Prenatal developmental origins of behavior and mental health: The influence of maternal stress in pregnancy. Vol. 117, Neuroscience and Biobehavioral Reviews. Elsevier Ltd; 2020. p. 26–64.

15. Moher D, Liberati A, Tetzlaff J, Altman DG, Grp P. Preferred Reporting Items for Systematic Reviews and Meta-Analyses: The PRISMA Statement (Reprinted from Annals of Internal Medicine). Phys Ther. 2009;89(9):873–80.

16. Jadad AR, Moore RA, Carroll D, Jenkinson C, Reynolds DJM, Gavaghan DJ, et al. Assessing the quality of reports of randomized clinical trials: Is blinding necessary? Control Clin Trials. 1996;17(1):1–12.

17. Verhagen AP, De Vet HCW, De Bie RA, Kessels AGH, Boers M, Bouter LM, et al. The Delphi list: A criteria list for quality assessment of randomized clinical trials for conducting systematic reviews developed by Delphi consensus. J Clin Epidemiol. 1998;51(12):1235–41.

18. Higgins JPT, Altman DG, Gøtzsche PC, Jüni P, Moher D, Oxman AD, et al. The Cochrane Collaboration’s tool for assessing risk of bias in randomised trials. BMJ. 2011;343(7829).

19. Beevi Z, Low WY, Hassan J. Impact of hypnosis intervention in alleviating psychological and physical symptoms during pregnancy. Am J Clin Hypn. 2016;58(4):368–82.

20. Davis K, Goodman SH, Leiferman J, Taylor M, Dimidjian S. A randomized controlled trial of yoga for pregnant women with symptoms of depression and anxiety. Complement Ther Clin Pract. 2015;21(3):166–72.

21. Chang MY, Chen CH, Huang KF. Effects of music therapy on psychological health of women during pregnancy. J Clin Nurs. 2008;17(19):2580–7.

22. Pan WL, Chang CW, Chen SM, Gau ML. Assessing the effectiveness of mindfulness-based programs on mental health during pregnancy and early motherhood - A randomized control trial. BMC Pregnancy Childbirth. 2019;19(1):1–8.

23. Rajeswari S, Sanjeevareddy N. Efficacy of progressive muscle relaxation on pregnancy outcome among anxious indian primi mothers. Iran J Nurs Midwifery Res. 2020;25(1):23–30.

24. Chang HC, Yu CH, Chen SY, Chen CH. The effects of music listening on psychosocial stress and maternal-fetal attachment during pregnancy. Complement Ther Med. 2015;23(4):509–15.

25. Liu YH, Lee CCS, Yu CH, Chen CH. Effects of music listening on stress, anxiety, and sleep quality for sleep-disturbed pregnant women. Women Heal. 2016;56(3):296–311.

26. Muthukrishnan S, Jain R, Kohli S, Batra S. Effect of mindfulness meditation on perceived stress scores and autonomic function tests of pregnant Indian women. J Clin Diagnostic Res. 2016;10(4):CC05–8.

27. Tragea C, Chrousos GP, Alexopoulos EC, Darviri C. A randomized controlled trial of the effects of a stress management programme during pregnancy. Complement Ther Med. 2014;22(2):203–11.

28. Bastani F, Hidarnia A, Kazemnejad A, Vafaei M, Kashanian M. A randomized controlled trial of the effects of applied pelaxation training on reducing anxiety and perceived stress in pregnant women. J Midwifery Women’s Heal. 2005;50(4).

29. Satyapriya M, Nagendra HR, Nagarathna R, Padmalatha V. Effect of integrated yoga on stress and heart rate variability in pregnant women. Int J Gynecol Obstet. 2009;104(3):218–22.

30. Newham JJ, Wittkowski A, Hurley J, Aplin JD, Westwood M. Effects of antenatal yoga on maternal anxiety and depression: A randomized controlled trial. Depress Anxiety. 2014;31(8):631–40.

31. Yang M, Li L, Zhu H, Alexander IM, Liu S, Zhou W, et al. To relieve anxiety in Pregnant women on bedrest: A randomized Trial, controlled trial. MCN Am J Matern Nurs. 2009;34(5):316–23.

32. Simavli S, Gumus I, Kaygusuz I, Yildirim M, Usluogullari B, Kafali H. Effect of music on labor pain relief, anxiety level and postpartum analgesic requirement: A randomized controlled clinical trial. Gynecol Obstet Invest. 2014;78(4):244–50.

33. Liu YH, Chang MY, Chen CH. Effects of music therapy on labour pain and anxiety in Taiwanese first-time mothers. J Clin Nurs. 2010;19(7–8):1065–72.

34. Bastani F, Hidarnia A, Montgomery KS, Aguilar-Vafaei ME, Kazemnejad A. Does relaxation education in anxious primigravid Iranian women influence adverse pregnancy outcomes? A randomized controlled trial. J Perinat Neonatal Nurs. 2006;20(2):138–46.

35. Chuntharapat S, Petpichetchian W, Hatthakit U. Yoga during pregnancy: Effects on maternal comfort, labor pain and birth outcomes. Complement Ther Clin Pract. 2008;14(2):105–15.

36. García González J, Ventura Miranda MI, Manchon García F, Pallarés Ruiz TI, Marin Gascón ML, Requena Mullor M, et al. Effects of prenatal music stimulation on fetal cardiac state, newborn anthropometric measurements and vital signs of pregnant women: A randomized controlled trial. Complement Ther Clin Pract. 2017;27:61–7.

37. Abu-Raya B, Michalski C, Sadarangani M, Lavoie PM. Maternal Immunological Adaptation During Normal Pregnancy. Front Immunol. 2020 Oct 7;11.

38. Sawle G V., Ramsay MM. The neurology of pregnancy. J Neurol Neurosurg Psychiatry. 1998;64(6):711–25.

39. Thornton CA. Immunology of pregnancy. Proc Nutr Soc. 2010 Aug;69(3):357–65.

40. Ohmori F, Shimizu S, Kagaya A. Exercise-induced blood flow in relation to muscle relaxation period. Dyn Med. 2007;6.

41. Kagaya A, Ogita F. Blood flow during muscle contraction and relaxation in rhythmic exercise at different intensities. Ann Physiol Anthropol. 1992;11(3):251–6.

42. Russo MA, Santarelli DM, O’Rourke D. The physiological effects of slow breathing in the healthy human. Breathe (Sheffield, England). 2017 Dec 1;13(4):298–309.

43. Field T. Postpartum depression effects on early interactions, parenting, and safety practices: A review. Vol. 33, Infant Behavior and Development. NIH Public Access; 2010. p. 1–6.

44. Chung TKH, Lau TK, Yip ASK, Chiu HFK, Lee DTS. Antepartum depressive symptomatology is associated with adverse obstetric and neonatal outcomes. Psychosom Med. 2001;63(5):830–4.

45. Dong Y, Yu JL. An overview of morbidity, mortality and long-term outcome of late preterm birth. World J Pediatr. 2011 Aug;7(3):199–204.

46. Sarkar P, Bergman K, O’Connor TG, Glover V. Maternal antenatal anxiety and amniotic fluid cortisol and testosterone: Possible implications for foetal programming. In: Journal of Neuroendocrinology. J Neuroendocrinol; 2008. p. 489–96.

47. Dale MTG, Bakketeig LS, Magnus P. Alcohol consumption among first-time mothers and the risk of preterm birth: A cohort study. Ann Epidemiol. 2016 Apr 1;26(4):275–82.

48. Marcus SM, Heringhausen JE. Depression in Childbearing Women: When Depression Complicates Pregnancy. Prim Care. 2009 Mar;36(1):151.

49. Van Den Bergh BRH, Mulder EJH, Mennes M, Glover V. Antenatal maternal anxiety and stress and the neurobehavioural development of the fetus and child: Links and possible mechanisms. A review. Neurosci Biobehav Rev. 2005;29(2):237–58.

50. O’Donnell K, O’Connor TG, Glover V. Prenatal stress and neurodevelopment of the child: Focus on the HPA axis and role of the placenta. Vol. 31, Developmental Neuroscience. Dev Neurosci; 2009. p. 285–92.

51. Wadhwa PD, Entringer S, Buss C, Lu MC. The Contribution of Maternal Stress to Preterm Birth: Issues and Considerations. Clin Perinatol. 2011 Sep;38(3):351.

52. Hobel CJ, Goldstein A, Barrett ES. Psychosocial stress and pregnancy outcome. Clin Obstet Gynecol. 2008 Jun;51(2):333–48.

53. Cardwell MS. Stress: pregnancy considerations. Obstet Gynecol Surv. 2013 Feb;68(2):119–29.

54. Witt WP, Cheng ER, Wisk LE, Litzelman K, Chatterjee D, Mandell K, et al. Maternal Stressful Life Events Prior to Conception and the Impact on Infant Birth Weight in the United States. Am J Public Health. 2014;104(Suppl 1):S81.

55. Davis EP, Glynn LM, Waffarn F, Sandman CA. Prenatal maternal stress programs infant stress regulation. J Child Psychol Psychiatry Allied Discip. 2011 Feb;52(2):119–29.

56. Nillni YI, Mehralizade A, Mayer L, Milanovic S. Treatment of depression, anxiety, and trauma-related disorders during the perinatal period: A systematic review. Clin Psychol Rev. 2018;66:136.

57. Dimidjian S, Goodman SH, Felder JN, Gallop R, Brown AP, Beck A. An open trial of mindfulness-based cognitive therapy for the prevention of perinatal depressive relapse/recurrence. Arch Womens Ment Health. 2015;18(1):85–94.

58. Fink NS, Urech C, Cavelti M, Alder J. Relaxation during pregnancy: What are the benefits for mother, fetus, and the newborn? A systematic review of the literature. J Perinat Neonatal Nurs. 2012;26(4):296–306.

59. Richter J, Bittner A, Petrowski K, Junge-Hoffmeister J, Bergmann S, Joraschky P, et al. Effects of an early intervention on perceived stress and diurnal cortisol in pregnant women with elevated stress, anxiety, and depressive symptomatology. J Psychosom Obstet Gynecol. 2012;33(4):162–70.

60. Alder J, Urech C, Fink N, Bitzer J, Hoesli I. Response to induced relaxation during pregnancy: Comparison of women with high versus low levels of anxiety. J Clin Psychol Med Settings. 2011;18(1):13–21.

